# Circulating lipids associate with future weight gain in individuals with an at-risk mental state and in first-episode psychosis

**DOI:** 10.1101/2020.01.30.20019711

**Authors:** Santosh Lamichhane, Alex M. Dickens, Partho Sen, Heikki Laurikainen, Jaana Suvisaari, Tuulia Hyötyläinen, Oliver Howes, Jarmo Hietala, Matej Orešič

**Affiliations:** Turku Bioscience Centre, University of Turku and Åbo Akademi University, Turku, Finland; Department of Psychiatry, University of Turku, FI-20520 Turku, Finland; Turku PET Centre, Turku University Hospital, FI-20521 Turku, Finland; Mental Health Unit, National Institute for Health and Welfare, Helsinki, Finland; Department of Chemistry, Örebro University, 70281 Örebro, Sweden; Department of Psychosis Studies, Institute of Psychiatry, Psychology & Neuroscience, King’s College London, London WC2R 2LS, UK; Psychiatric Imaging Group, MRC London Institute of Medical Sciences, Hammersmith Hospital, Imperial College London, London W12 0HS, UK; School of Medical Sciences, Örebro University, 70281 Örebro, Sweden

## Abstract

Patients with schizophrenia have a lower than average life span, largely due to the increased prevalence of cardiometabolic co-morbidities. Identification of individuals with psychotic disorders with a high risk of rapid weight gain, and the associated development of metabolic complications, is an unmet need as regards public health. Here, we applied mass spectrometry-based lipidomics in a prospective study comprising 48 controls (CTR), 44 first-episode psychosis (FEP) patients and 22 individuals at clinical-high-risk (CHR) for psychosis, from two study centers (Turku/Finland and London/UK). Baseline serum samples were analyzed by lipidomics, while body mass index (BMI) was assessed at baseline and after 12 months. We found that baseline triacylglycerols with low double bond counts and carbon numbers were positively associated with the change in BMI at follow-up. In addition, a molecular signature comprised of two triacylglycerols (TG(48:0) and TG(45:0)), was predictive of weight gain in individuals with a psychotic disorder, with an area under the receiver operating characteristic curve (AUROC) of 0.74 (95% CI: 0.60–0.85). When independently tested in the CHR group, this molecular signature predicted said weight change with AUROC = 0.73 (95% CI: 0.61–0.83). We conclude that molecular lipids may serve as a predictor of weight gain in psychotic disorders in at-risk individuals, and may thus provide a useful marker for identifying individuals who are most prone to developing cardiometabolic co-morbidities.

## Introduction

Psychotic disorders are associated with a life expectancy reduction of 15-20 years [1,2], mostly due to the high prevalence of cardiovascular disease, type 2 diabetes (T2DM) and metabolic syndrome [3-5]. Metabolic co-morbidities, including impaired glucose tolerance, weight gain and obesity often co-occur in first episode psychosis (FEP) patients [6-8], and this increases the risk of cardiovascular disease in these individuals [9,10]. Although unhealthy lifestyles and antipsychotic medication associate with the development of metabolic co-morbidities in psychosis patients, the underlying mechanisms remain poorly understood [3,11]. Drug-induced metabolic dysregulation appears heterogeneously [12,13], while metabolic co-morbidities can also occur in drug-naïve FEP patients [6,14].

Metabolomics, that is, a global study of small molecules (< 1500 Da) and their associated biochemical processes, is a powerful emerging tool in psychiatric research, enabling investigations of disease etiology and treatment responses from metabolic perspectives [8,15]. Lipidomics is a sub-field of metabolomics, which focuses on study of lipids. By applying a lipidomics approach, we have previously found that FEP patients who rapidly gain weight during follow-up have increased serum lipids at baseline; lipids which are also known to be associated with non-alcoholic fatty liver disease (NAFLD) and increased risk T2DM [8,16].

However, it is currently unclear if these lipids could be used for prediction of said weight gain and the associated metabolic co-morbidities in FEP patients. Here we report a lipidomics study in a prospective series of plasma samples from healthy controls (CTR), FEP patients (FEP), and individuals at clinical-high-risk (CHR) for psychosis. The aim was to investigate the ability of lipid profiles to identify FEP patients or CHR individuals, who are at the highest risk of rapid weight gain and occurrence of metabolic co-morbidities.

## Methods

### Study design and participants

We collected plasma samples from two cohorts of patients receiving psychiatric early intervention services in Turku, Finland or London, United Kingdom. Ethical approval was obtained from the respective study sites in Finland (ETMK 98/180/2013) and United Kingdom (14/LO/1289). Capacity for consent was assessed and informed written consent was obtained from all volunteers. In total, 114 non-fasting blood samples were collected for this study. This case-control study included 48 healthy controls (CTR group), 44 first-episode psychosis patients (FEP group) and 22 individuals who were at clinical-high-risk for psychosis (CHR group). Demographic characteristics of the study subjects are shown in **Table 1**.

**Table 1.** Clinical characteristics of study population. Abbreviations: CTR, healthy controls; FEP, first-episode psychosis group; CHR, clinical high-risk for psychosis group; SD, standard deviation; CPZE, Chlorpromazine equivalence.

|  | **CTR** | **FEP** | **CHR** |
| --- | --- | --- | --- |
| N (Total) | 48 | 44 | 22 |
| n (Turku, Finland) | 31 | 30 | 22 |
| n (London, UK) | 17 | 14 | N/A |
| Sex (Male, Female) | 31, 17 | 26, 18 | 11, 11 |
| BMI (± SD) | 24.50 (3.85) | 24.35 (4.26) | 25.56 (5.72) |
| Ethnicity (EU, non-EU, NA) | 36, 12, -- | 37, 6, 1 | 22 |
| GAF score (± SD) | 92.40 (3.80) | 47.80 (16.37) | 56.00 (9.84) |
| PANSS TOT (± SD) | 30.48 (0.97) | 70.07 (24.40) | 54.33 (12.65) |
| Antipsychotic CPZE (± SD) | | | |
| Turku, Finland<br>(n = 23 FEP, n = 9 CHR) | N/A | 221.83 (± 115.04) | 207.91(± 102.23) |
| London, UK<br>(n = 13 FEP) | N/A | 622.63 (± 650.94) | N/A |

\* Study populations are from Turku/Finland and London/UK

FEP patients met the following inclusion criteria: (i) DSM-IV diagnosis of a psychotic disorder, determined by the Structured Interview for Prodromal Syndromes (SCID)-I/P; (ii) illness duration of less than 3 years. In the Turku/Finland study, FEP volunteers were taking antipsychotic medication and had diagnoses of affective or non-affective psychosis. In the London/UK, FEP arm of the study, volunteers were medication-free from all pharmacological treatments for at least 6 months and had diagnoses of schizophrenia or schizoaffective disorder. In the London/UK cohort, FEP volunteers were recruited from first episode teams covering central south London (total population approximately 1.5 million people). The state-funded health service in the UK means that these teams receive all referrals with a first-episode of psychosis within the catchment area. We recruited patients who were medication-free for at least 6 months and had diagnoses of schizophrenia or schizoaffective disorder. Healthy volunteers had no current/lifetime history of an Axis-I disorder as determined by the SCID-I/P and were matched by age (age +/- 3 years).

CHR patients were identified from the clinical population of psychiatric services using SCID interviews [17]. Patients with either brief, intermittent psychotic symptom syndrome, attenuated positive symptom syndrome or genetic risk and deterioration syndrome were classified as clinical high risk for psychosis patients [17].

The study setting for the THL/Finland dataset, which was used as an additional dataset to build the statistical model, was described in detail previously [18].

### Analysis of molecular lipids

A total of 114 plasma samples were randomized and extracted using a modified version of the Folch procedure [19]. Promptly before extraction, 10 µL of 0.9% NaCl and 120 µL of CHCl3: MeOH (2:1, v/v) containing 2.5 µg mL^-1^ internal standard solution (for quality control (QC) and normalization purposes) were added to 10 µL of each plasma sample. The standard solution contained the following compounds: 1,2-diheptadecanoyl-sn-glycero-3-phosphoethanolamine (PE(17:0/17:0)), N-heptadecanoyl-D-erythrosphingosylphosphorylcholine (SM(d18:1/17:0)), N-heptadecanoyl-D-erythro-sphingosine (Cer(d18:1/17:0)), 1,2-diheptadecanoyl-sn-glycero-3-phosphocholine (PC(17:0/17:0)), 1-heptadecanoyl-2-hydroxy-sn-glycero-3-phosphocholine (LPC(17:0)) and 1-palmitoyl-d31-2-oleoyl-sn-glycero-3-phosphocholine (PC(16:0/d31/18:1 that were purchased from Avanti Polar Lipids, Inc. (Alabaster, AL, USA), as well as 3β-Hydroxy-5-cholestene 3-heptadecanoate (CE17:0) and tripalmitin-triheptadecanoylglycerol (TG(17:0/17:0/17:0)) (Larodan AB, Solna, Sweden). The samples were vortexed and incubated on ice for 30 min after which they were centrifuged (9400 × g, 3 min, 4 °C). 60 µL from the lower layer of each sample was then transferred to a glass vial with an insert, and 60 µL of CHCl3: MeOH (2:1, v/v) was added to each sample. The samples were re-randomized and stored at −80 °C until analysis. Calibration curves using 1-hexadecyl-2-(9Z-octadecenoyl)-sn-glycero-3-phosphocholine (PC(16:0/18:1(9Z))), 1-(1Z-octadecenyl)-2-(9Z-octadecenoyl)-sn-glycero-3-phosphocholine (PC(16:0/16:0)), 1,2-dihexadecanoyl-sn-glycero-3-phosphocholine (PC(18:0/18:0), 1-octadecanoyl-sn-glycero-3-phosphocholine (LPC(18:0)), 1-(11Z-octadecadienoyl)-sn-glycero-3-phosphocholine (LPC(18:1)), 1-(9Z-octadecenoyl)-2-hexadecanoyl-sn-glycero-3-phosphoethanolamine (PE (16:0/18:1)), (2-aminoethoxy)[(2R)-3-hydroxy-2-[(11Z)-octadec-11-enoyloxy]propoxy]phosphinic acid (LysoPE (18:1)), N-(9Z-octadecenoyl)-sphinganine (Cer (d18:0/18:1(9Z))), 1-hexadecyl-2-(9Z-octadecenoyl)-sn-glycero-3-phosphoethanolamine (PE (16:0/18:1)) (Avanti Polar Lipids, Inc.), 1-Palmitoyl-2-Hydroxy-sn-Glycero-3-Phosphatidylcholine (LPC(16:0)) and 1,2,3 trihexadecanoalglycerol (TG16:0/16:0/16:0), 1,2,3-trioctadecanoylglycerol (TG(18:0/18:0/18:0)) and ChoE (18:0), 3β-Hydroxy-5-cholestene 3-linoleate (ChoE(18:2)) (Larodan AB, Solna, Sweden), were prepared at the following concentrations: 100, 500, 1000, 1500, 2000 and 2500 ng mL^−1^ (in CHCl3:MeOH, 2:1, v/v) including 1250 ng mL^-1^ of each internal standard. The samples were analyzed using an established ultra-high-performance liquid chromatography quadrupole time-of-flight mass spectrometry method (UHPLC-QTOFMS). The UHPLC system used in this work was a 1290 Infinity system from Agilent Technologies (Santa Clara, CA, USA). The system was equipped with a multi sampler (maintained at 10 °C), a quaternary solvent manager and a column thermostat (maintained 7 at 50 °C). Separations were performed on an ACQUITY UPLC® BEH C18 column (2.1 mm × 100 mm, particle size 1.7 µm) by Waters (Milford, USA). The mass spectrometer coupled to the UHPLC was a 6545 quadrupole time of flight (QTOF) from Agilent Technologies interfaced with a dual jet stream electrospray ion (dual ESI) source. All analyses were performed in positive ion mode and MassHunter B.06.01 (Agilent Technologies) was used for all data acquisition. Quality control was performed throughout the sample run by including blanks, pure standard samples, extracted standard samples and control plasma samples. An aliquot of each sample was collected and pooled and used as quality control sample, together with NIST SRM 1950 reference plasma sample [20], an in-house pooled serum sample.

Relative standard deviations (% RSDs) for lipid internal standards representing each lipid class in the samples (raw variation) was below 11%. The lipid concentrations in the pooled control samples were, on average, 16.4% (KCL) and 11.4% (UTU). This shows that the method is reliable and reproducible throughout the sample set.

The identification was carried out in pooled serum sample, and with this information, an in-house database was created with m/z and retention time for each lipid. Identification of lipids was carried out by combining MS (and retention time), MS/MS information, and a search of the LIPID MAPS spectral database [21], and in some cases by using authentic lipid standards. MS/MS data were acquired in both negative and positive ion modes in order to maximize identification coverage. The confirmation of a lipid’s structure requires the identification of hydrocarbon chains bound to its polar moieties, and this was possible in some cases.

### Data pre-processing

Mass spectrometry (MS) data processing was performed using the open-source software, MZmine 2.18 [22]. The following steps were applied in the processing: (1) Crop filtering with a m/z range of 350 – 1200 m/z and a RT range of 2.0 to 15.0 min, (2) Mass detection with a noise level of 1000, (3) Chromatogram builder with a min time span of 0.08 min, min height of 1200 and a m/z tolerance of 0.006 m/z or 10.0 ppm, (4) Chromatogram deconvolution using the local minimum search algorithm with a 70% chromatographic threshold, 0.05 min minimum RT range, 5% minimum relative height, 1200 minimum absolute height, a minimum ration of peak top/edge of 1.2 and a peak duration range of 0.08 - 5.0, (5) Isotopic peak grouper with a m/z tolerance of 5.0 ppm, RT tolerance of 0.05 min, maximum charge of 2 and with the most intense isotope set as the representative isotope, (6) Peak list row filter keeping only peaks with a minimum of 10 peaks in a row, (7) Join aligner with a m/z tolerance of 0.009 or 10.0 ppm and a weight for of 2, a RT tolerance of 0.1 min and a weight of 1 and with no requirement of charge state or ID and no comparison of isotope pattern, (8) Peak list row filter with a minimum of 53 peak in a row (10% of the samples), (9) Gap-filling using the same RT and m/z range gap filler algorithm with an m/z tolerance of 0.009 m/z or 11.0 ppm, (10) Identification of lipids using a custom database search with an m/z tolerance of 0.009 m/z or 10.0 ppm and a RT tolerance of 0.1 min, (11) Normalization using internal class-specific standards (PE (17:0/17:0), SM (d18:1/17:0), Cer (d18:1/17:0), LPC (17:0), TG (17:0/17:0/17:0) and PC (16:0/d30/18:1)) for identified lipids and closest ISTD for the unknown lipids, followed by calculation of the estimated concentrations based on lipid-class calibration curves, (12) Imputation of missing values was calculated as half of the lipid’s minimum observed value.

### Data analysis

Mann-Whitney U test was applied to compare the difference in weight gain between the study groups (*e*.*g*., CTR *vs*. FEP), and performed using GraphPad Prism v. 7.04 (GraphPad Software Inc., San Diego, CA). In order to visualize the changes in BMI between the study groups, we created a violin plot using the ggplot2 package (version 3.2.1) in the R statistical software [23]. Spearman correlation coefficients were calculated using the statistical toolbox in MATLAB 2017b (Mathworks Inc., Natick, MA) and p-values < 0.05 (two-tailed) were considered significant for these correlations. All statistical analyses involving lipid concentrations were performed on log2-transformed data. The mclust R package (version 5.4.1) was used to build lipid clusters from the lipidomics dataset. Mclust allows modelling of data as a Gaussian finite mixtures and attempts to fit various model types and assesses their performance using the Bayesian Information Criterion (BIC). The highest BIC achieved by mclust form the lipidomics dataset in control subjects was used to determine both the model type and the number of clusters into which the variables should be divided.

Logistic ridge regression (LR) models were developed to predict and stratify weight gain in the FEP patients. The matched triacylglycerols (TGs) with a regression coefficient (r ≥ 0.4) in Turku/Finland and London/UK cohorts, between high *vs*. low weight gain subjects (*i*.*e*., change in the BMIs binarized around the median), were used either singly or in combination for LR modelling. A recursive feature elimination scheme was implemented for the optimal selection of the lipids. The lipids in the LR models were incorporated or removed in an iterative manner, starting with all nine TGs. The models were adjusted for sex and assessed by area under the receiver operating characteristic (ROC) curves (AUROCs). The mean AUROC of a model was estimated by bootstrapping, *i*.*e*., 1000 times re-sampling without replacement and partitioning (70% training, 30% test sets) of the lipidomic dataset using ‘*createDataPartition’* function coded in the*’caret’ (v. 6*.*0*.*84)* R package. The model with the highest mean AUROC was considered to be the best model which was assessed by their ROC curves using *‘pROC 1*.*15*.*3’* R package. Regularized ridge models in *‘cv*.*glmnet’* requires a hyper-parameter ‘λ’. Here, the λ_minimum_ that corresponds to the minimum cross-validation (CV) error was determined by 10-fold CV. The LR model with the highest mean AUROC was named FEP-LR model. This model was also used to predict weight gain (change in BMI from the baseline) an independent dataset (the CHR subjects).

## Results

### Lipidome in first episode psychosis patients

We measured circulating molecular lipids by UHPLC-QTOFMS from the three study groups (**Fig. 1**), together comprising 48 healthy controls (CTR), 44 FEP patients and 22 CHR individuals, from two study centers (Turku, Finland and London, UK), at baseline as well as at one-year follow-up (CTR, n=21; FEP n=13; CHR, n=9). Demographic characteristics of the three study groups are shown in **Table 1**. The lipidomics dataset included in data analysis comprised 265 identified lipids from the following lipid classes: cholesterol esters (CE), ceramides (Cer), lysophosphatidylcholines (LPC), phosphatidylcholines (PC), phosphatidylethanolamines (PE), sphingomyelins (SM) and triacylglycerols (TG).

**Figure 1.**
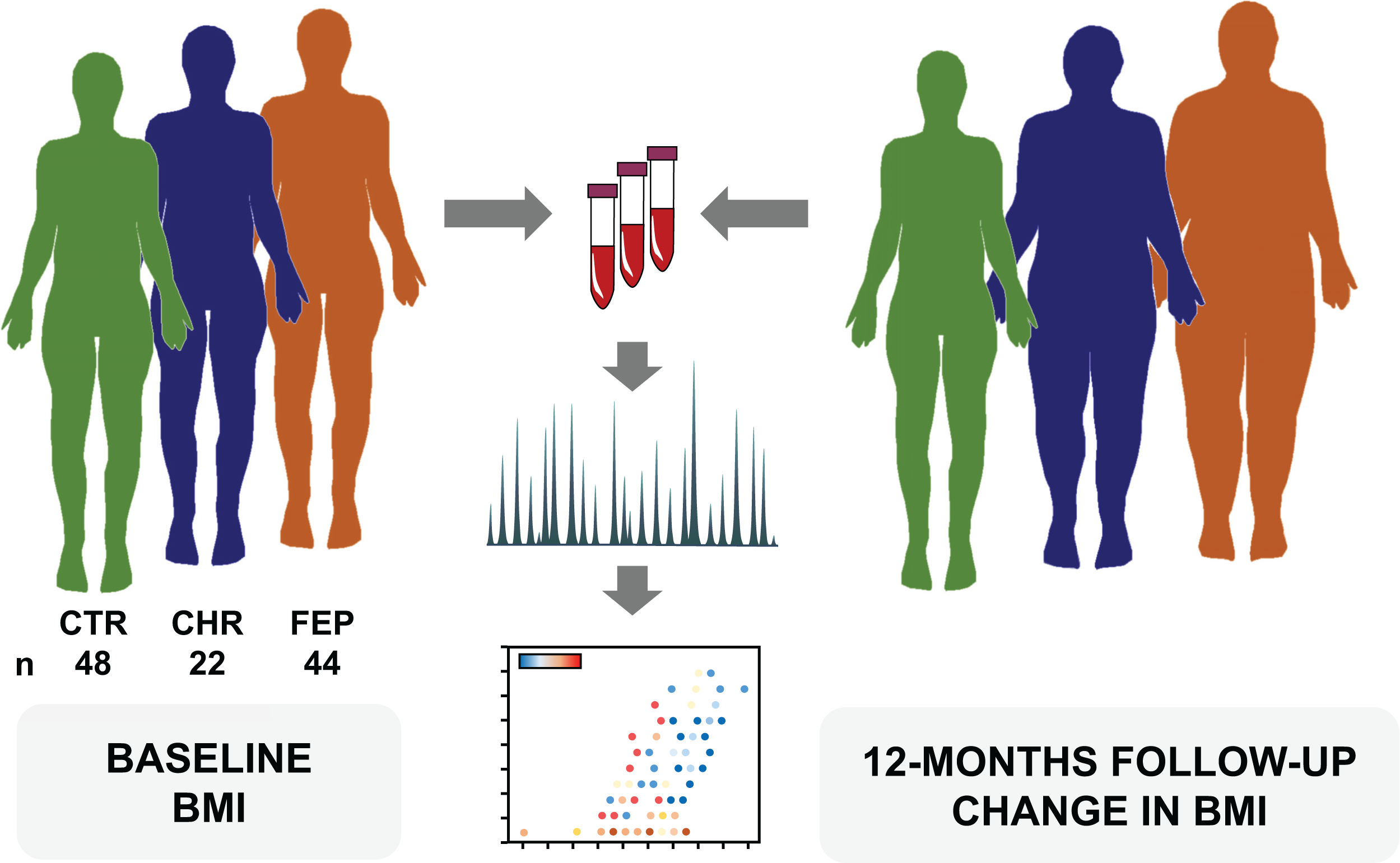
Study setting. Lipidomics was applied to analyze baseline samples from 48 healthy controls (CTR), 44 first-episode psychosis patients (FEP) and 22 individuals at clinical-high-risk for psychosis (VHR), from two study centers (Turku, Finland and London, UK). Body mass index (BMI) and other metabolic measures were assessed at baseline and at 12-month follow-up.

In order to summarize the data, we first performed clustering using the Gaussian mixture models [24], reducing the data into 22 distinct lipid clusters (**Supplementary Table 1**). As expected, the lipids were clustered according to the main functional lipid classes. For example, PCs and SMs predominated in lipid clusters (LCs) LC3 and LC6, while LPCs and Cers had distinct clusters (LC4 and LC5, respectively). These clusters also revealed sub-grouping according to the acyl chain carbon number and double-bond count in TGs (LC13, LC14).

### Associations between lipidome and weight gain

We then examined the differences in weight gain between the study groups (CTR *vs*. FEP, CTR *vs*. CHR, and CHR *vs*. FEP). FEP patients gained weight when compared to the CTR group (**Fig 2a**; p = 0.004). No significant differences were observed when comparing CHR *vs*. FEP and CTR *vs*. CHR (p = 0.3851 for CHR *vs*. FEP and p = 0.0561 for CHR *vs*. CTR).

**Figure 2.**
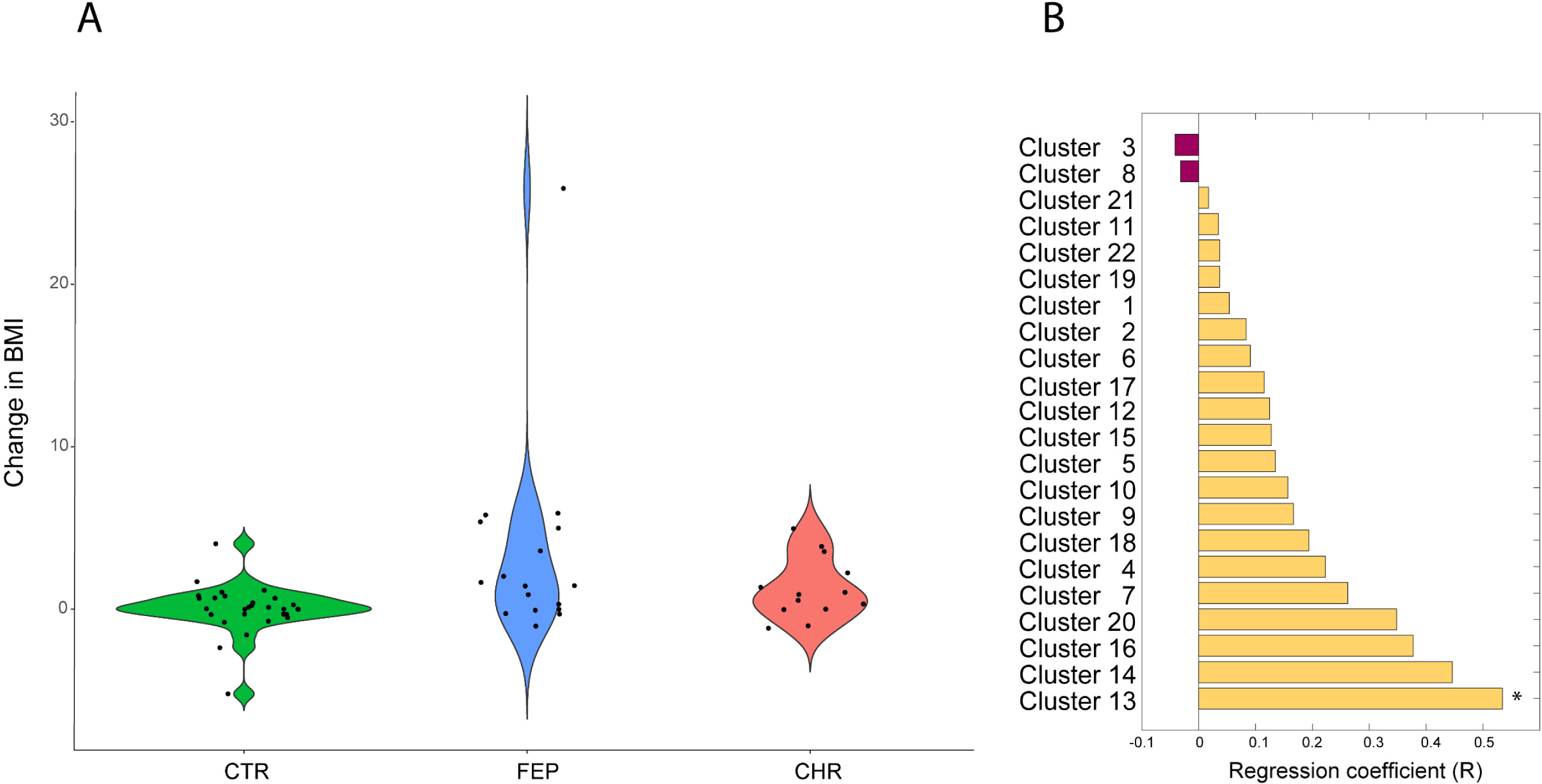
Associations between lipidome and weight gain. a) Difference in BMI change (12-month follow-up *vs*. baseline) between the study groups (CTR *vs*. FEP, CTR *vs*. CHR, and CHR *vs*. FEP). b) Association between baseline lipid clusters and weight gain in FEP group. *p<0.05.

Next, we analyzed the association between the mean levels of the lipids in the baseline lipid clusters and weight gain in CTR and FEP groups. Among the 22 LCs, the baseline level of cluster LC13 was associated with changes in BMI in the FEP group after 12-month follow-up time (Spearman r = 0.53, p =0.0291). The cluster LC13 contains TGs with low double-bond count, indicating that the change of BMI in FEP patients was specifically associated with a structurally-distinct subgroup of lipids. Interestingly, we observed trends of positive association (r > 0.3) between weight gain and other lipid clusters containing mainly TGs (L14, L16, L20; **Fig 2b**). Thus, we sought to determine the association between baseline TG composition and change of BMI (12-month follow-up vs. baseline) at the molecular lipid level. The baseline levels of TGs with low carbon number and double bond count showed positive associations with the change in BMI among the FEP patients (**Fig. 3b**), while the association in the CTR group remained weak (**Fig. 3a**). Nine of 109 TGs at baseline, including TG(47:0), TG(47:1), TG(48:0), TG(48:0) TG(48:1), TG(48:1), TG(49:0), TG(14:0/16:0/18:1), and TG(16:0/16:0/16:0), were significantly associated with the change in BMI (p < 0.05, **Supplementary Table 2**). Similarly, we performed correlation analysis between changes in BMI and baseline TG composition in CHR individuals. 32 out of 109 TGs remained correlated with the change in the BMI (p < 0.05, **Supplementary Table 3**). Baseline TGs with low carbon number and double bond count showed strong positive association with the change in BMI (**Fig. 3c**).

**Figure 3.**
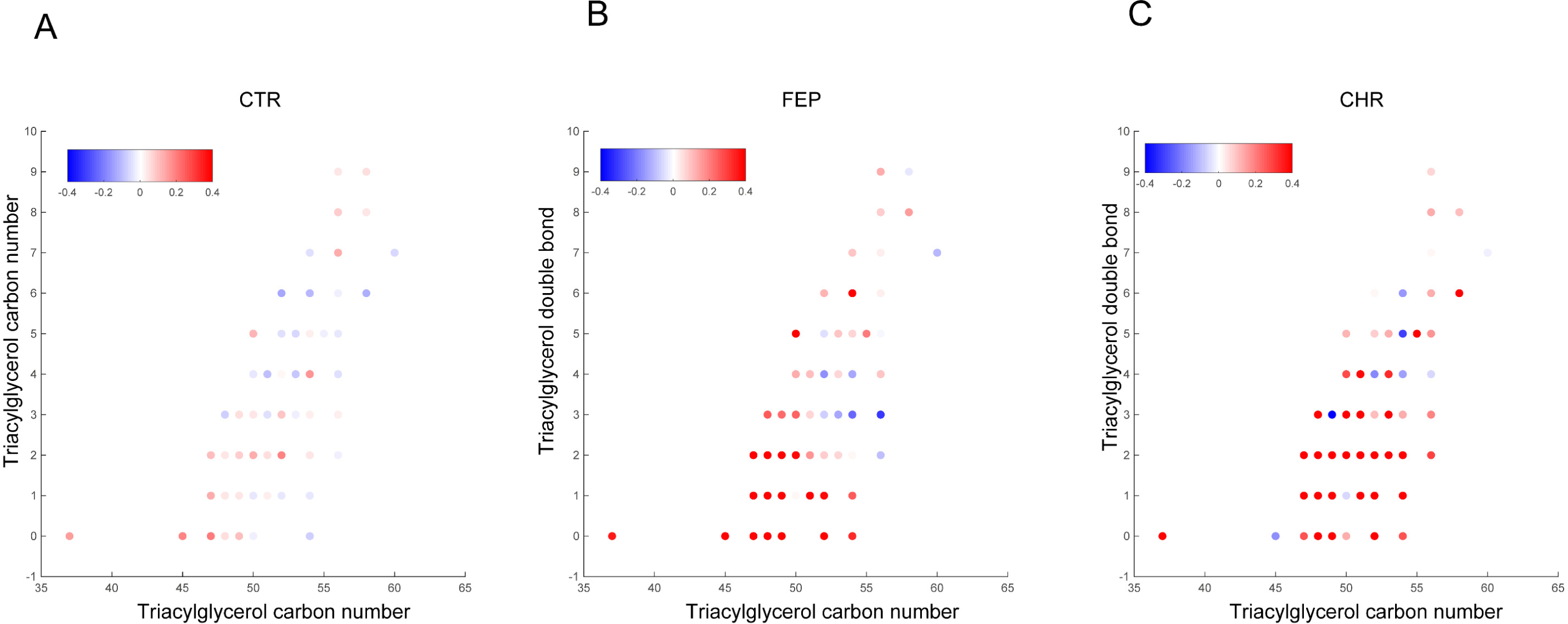
Correlation of individual TGs with change in BMI (12-month follow-up *vs*. baseline). The x-axis is the acyl carbon number and y-axis is the acyl double bond count. a) CTR, b) FEP and c) CHR. The spearman correlation coefficient (R) is used for the color code.

### Prediction of weight gain in FEP patients and CHR individuals by circulating lipids

Next, we sought to determine if baseline TG concentrations predicted the risk of weight gain in FEP patients, utilizing the regularized logistic regression (LR) model. We examined the predictor model combining the data from three centers including Turku/Finland, London/UK, and the matched TGs from the THL/Finland dataset. The matched TGs with regression coefficient (r ≥ 0.4) in Turku/Finland and London/UK cohorts were used as input to build the LR models between the high and low weight gain groups (binarized at their median change of BMIs from the baseline, see **Methods**) in FEP cases. The recursive scheme for feature selection and model reduction showed that TG (48:0) together with TG (45:0) were the best predictors for high change in BMI, with AUROC=0.74 (**Fig. 4a**, 95% confidence interval, CI: 0.60 – 0.85).

**Figure 4.**
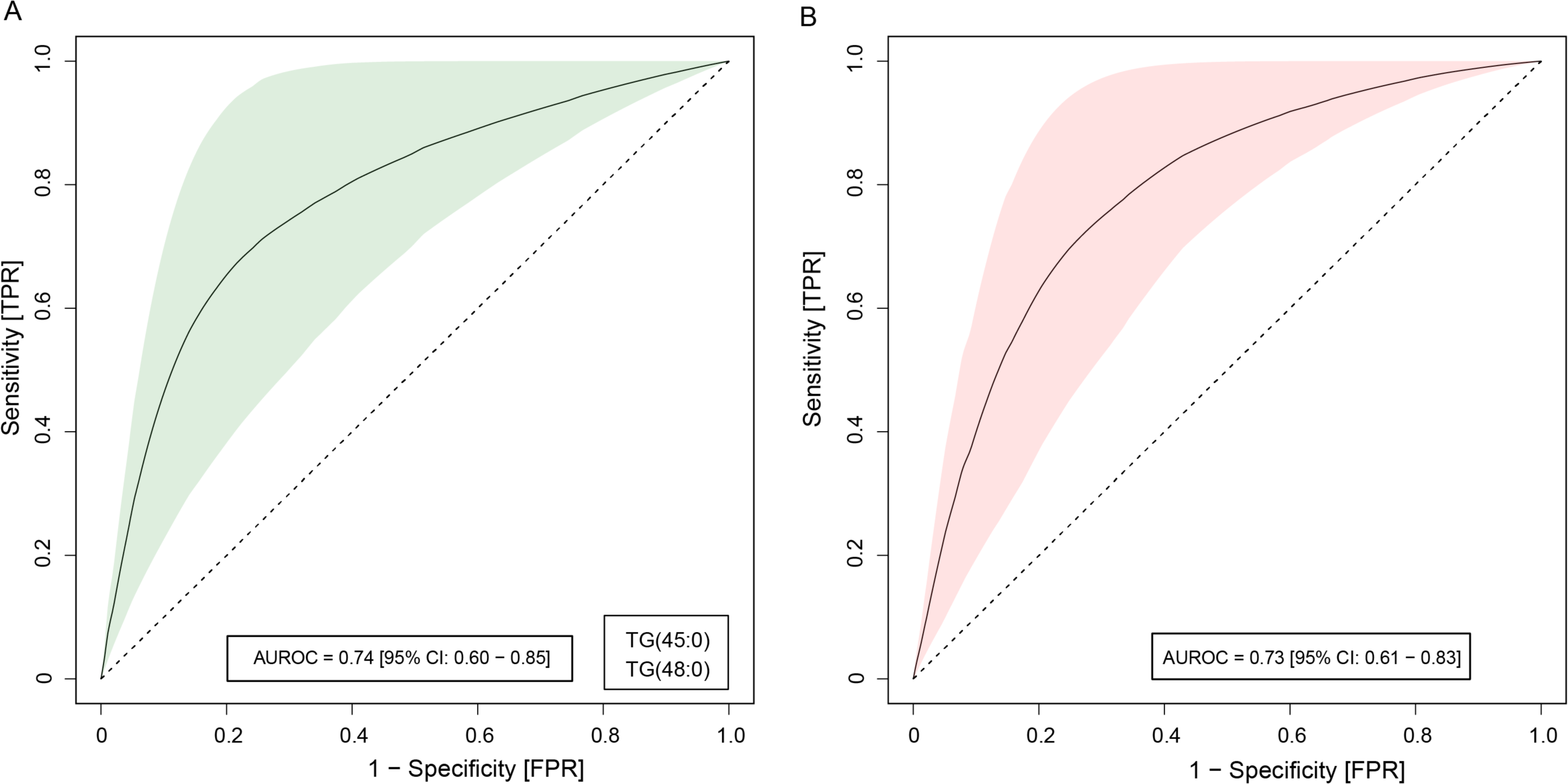
Predictive models of weight gain (BMI change in the 12-month follow-up) in the FEP and CHR group. Logistic ridge regression (LR) models showing triacylglycerols (TGs) as predictive markers to stratify patient groups into high and low BMI changes. a) Receiver-operating characteristic (ROC) curves, showing the performance of the LR models with highest mean AUROCs in the FEP patients, discriminating high *vs*. low BMI changes in 12-month follow-up. The light green shaded area denotes the 95% confidence intervals (CI), as calculated by using bootstrapping. b) ROC curves showing the prediction performance of the FEP-LR models with highest mean AUROCs in the CHR individuals, discriminating high *vs*. low BMI changes in 12-month follow-up.

We then independently tested the potential of the FEP-LR model to predict weight gain (change in BMI) in CHR individuals. The FEP-LR model was indeed able to predict the outcome with AUROC=0.73 (**Fig. 4b**, 95% CI: 0.61 – 0.83).

## Discussion

Our study demonstrates that circulating lipids can predict risk of future weight gain in FEP patients and, as a novel finding, also in CHR individuals. We found that plasma lipids, specifically TGs, may form a useful molecular signature for the identification of individuals who are vulnerable to rapid weight gain. This finding is in line with and builds on our previous study, which showed that weight gain in FEP patients is associated with elevated TGs containing low acyl carbon numbers and double bond counts, independently of obesity at baseline [16,25].

TGs with low double bond count and carbon number, which are, in part, generated by *de novo* lipogenesis [26,27], are known to be elevated in non-alcoholic fatty liver disease (NAFLD) [28-30] and associated with an increased risk of T2DM [31,32]. Our findings thus strongly suggest that the FEP patients who go on to gain weight in the future are those who have elevated levels of liver fat.

Weight gain and metabolic co-morbidities are typically evident in antipsychotic drug-naïve FEP patients [11,33]. However, there is considerable variability in weight gain and lipid changes among the FEP individuals with respect to antipsychotic drugs [34-36]. Due to the relatively small sample size in the present study, we could not systematically examine the impact of specific antipsychotic drugs on weight gain, and their association with the baseline lipid levels. However, earlier analyses suggest that the NAFLD lipid signature associates with weight gain, independent of antipsychotic medication [16]. In line with this, and as a novel finding, we have here also demonstrated that the same lipid signature, predictive of weight-gain in FEP patients, is also predictive of weight gain in CHR individuals. This suggests that specific lipid disturbances in early psychosis may also contribute to the development of metabolic co-morbidities, potentially independent of antipsychotic medication. Since a fraction of CHR individuals in our study received low-dose antipsychotic medication, one also cannot exclude the possibility that the metabolic consequences in some CHR individuals may have been influenced by the use of antipsychotics [37].

The specific mechanisms linking psychosis, NAFLD, and increased risk of metabolic co-morbidities, are currently unknown. One plausible, yet currently speculative link, is the endocannabinoid system (ECS). Our positron emission tomography (PET) imaging data suggest that the brain ECS is dysregulated in FEP, including in drug-naïve patients [38]. Brain CB1R availability, as measured by PET imaging, associates with changes in peripheral endocannabinoid levels [39]. Furthermore, there is a large body of literature suggesting that the ECS modulates energy intake [40], and that the development of NAFLD is promoted by peripheral activation of the ECS [41]. More studies are clearly needed if one is to elucidate the hypothetical role of ECS as a link between psychosis and the development of metabolic co-morbidities.

Taken together, our study independently confirms that the lipidomic signature of NAFLD may serve as a predictor of future weight gain in FEP patients as well as in CHR individuals. This lipid signature may be used for the identification of at-risk individuals and patients who are at increased risk for development of metabolic co-morbidities in psychosis. Such knowledge may be useful in targeting primary prevention of metabolic co-morbidities and the identification of optimal treatment strategies for each patient.

## Data Availability

The lipidomics dataset and the relevant clinical metadata generated in this study were submitted to MetaboLights database, under accession number MTBLS1467.

## Acknowledgements

The authors thank to Cecilia Karlsson for assistance with lipidomics analysis and Aidan McGlinchey for editing. We also thank the following METSY project investigators: Raimo K. R. Salokangas, Tuula Ilonen, Päivi Jalo, Akseli Mäkelä, Tiina From, Janina Paju, Anna Toivonen, Reetta-Liina Armio, Mirka Kolkka, Maija Walta, Max Karukivi, Juha Mäkelä, Maria Tikka, Olof Solin, Merja Haaparanta-Solin, Aidan McGlinchey, Juha Pajula, Mark van Gils, Juha M. Kortelainen, Carmen Moreno, Joost Janssen, Javier Santonja, Covadonga M. Diaz-Caneja, Miriam Ayora Rodriguez, Celso Arango, Alberto Rodriguez-Quiroga, Fabian Hernández-Álvarez, Jose L. Ayuso-Mateos, Roberto Rodriguez-Jimenez, Angela Ibañez, Jaana Suvisaari, Maija Lindgren, Teemu Mäntylä, Tuula Kieseppä, Outi Mantere, Eva Rikandi, Tuukka T. Raij, Dieter Maier, Elisabeth Frank, Markus Butz-Ostendorf.

## Author contributions

M.O., J.H., and O.H. initiated, designed, and supervised the study. H.L. and METSY investigators recruited the subjects, performed the clinical interviews and collected the blood samples. A.D. and T.H. acquired serum lipidomics data. P.S., A.D., and S.L. analyzed the data. S.L. and M.O. wrote the first draft of the manuscript. All authors approved the final version.

## References

1 McGrath JJ, Saha S. Thought experiments on the incidence and prevalence of schizophrenia “under the influence” of cannabis. Addiction. 2007;102(4):514-5; discussion 16-8.

2 Brown S. Excess mortality of schizophrenia. A meta-analysis. Br J Psychiatry. 1997;171:502–8.

3 Henderson DC, Vincenzi B, Andrea NV, Ulloa M, Copeland PM. Pathophysiological mechanisms of increased cardiometabolic risk in people with schizophrenia and other severe mental illnesses. Lancet Psychiatry. 2015;2(5):452–64.

4 Mukherjee S, Schnur DB, Reddy R. Family history of type 2 diabetes in schizophrenic patients. Lancet. 1989;1(8636):495.

5 Ringen PA, Engh JA, Birkenaes AB, Dieset I, Andreassen OA. Increased mortality in schizophrenia due to cardiovascular disease - a non-systematic review of epidemiology, possible causes, and interventions. Front Psychiatry. 2014;5:137.

6 Pillinger T, Beck K, Gobjila C, Donocik JG, Jauhar S, Howes OD. Impaired Glucose Homeostasis in First-Episode Schizophrenia: A Systematic Review and Meta-analysis. JAMA Psychiatry. 2017;74(3):261–69.

7 Oresic M. Obesity and psychotic disorders: uncovering common mechanisms through metabolomics. Dis Model Mech. 2012;5(5):614–20.

8 Oresic M, Tang J, Seppanen-Laakso T, Mattila I, Saarni SE, Saarni SI, et al. Metabolome in schizophrenia and other psychotic disorders: a general population-based study. Genome Med. 2011;3(3):19.

9 Vancampfort D, Stubbs B, Mitchell AJ, De Hert M, Wampers M, Ward PB, et al. Risk of metabolic syndrome and its components in people with schizophrenia and related psychotic disorders, bipolar disorder and major depressive disorder: a systematic review and meta-analysis. World Psychiatry. 2015;14(3):339–47.

10 Emul M, Kalelioglu T. Etiology of cardiovascular disease in patients with schizophrenia: current perspectives. Neuropsychiatr Dis Treat. 2015;11:2493–503.

11 Bak M, Fransen A, Janssen J, van Os J, Drukker M. Almost all antipsychotics result in weight gain: a meta-analysis. PLoS One. 2014;9(4):e94112.

12 Le Hellard S, Theisen FM, Haberhausen M, Raeder MB, Ferno J, Gebhardt S, et al. Association between the insulin-induced gene 2 (INSIG2) and weight gain in a German sample of antipsychotic-treated schizophrenic patients: perturbation of SREBP-controlled lipogenesis in drug-related metabolic adverse effects? Mol Psychiatry. 2009;14(3):308–17.

13 MacKenzie NE, Kowalchuk C, Agarwal SM, Costa-Dookhan KA, Caravaggio F, Gerretsen P, et al. Antipsychotics, Metabolic Adverse Effects, and Cognitive Function in Schizophrenia. Front Psychiatry. 2018;9:622.

14 Tarricone I, Ferrari Gozzi B, Serretti A, Grieco D, Berardi D. Weight gain in antipsychotic-naive patients: a review and meta-analysis. Psychol Med. 2010;40(2):187–200.

15 Oresic M, Seppanen-Laakso T, Sun D, Tang J, Therman S, Viehman R, et al. Phospholipids and insulin resistance in psychosis: a lipidomics study of twin pairs discordant for schizophrenia. Genome Med. 2012;4(1):1.

16 Suvitaival T, Mantere O, Kieseppa T, Mattila I, Poho P, Hyotylainen T, et al. Serum metabolite profile associates with the development of metabolic co-morbidities in first-episode psychosis. Transl Psychiatry. 2016;6(11):e951.

17 Miller TJ, McGlashan TH, Rosen JL, Cadenhead K, Cannon T, Ventura J, et al. Prodromal assessment with the structured interview for prodromal syndromes and the scale of prodromal symptoms: predictive validity, interrater reliability, and training to reliability. Schizophr Bull. 2003;29(4):703–15.

18 Keinanen J, Mantere O, Kieseppa T, Mantyla T, Torniainen M, Lindgren M, et al. Early insulin resistance predicts weight gain and waist circumference increase in first-episode psychosis--A one year follow-up study. Schizophr Res. 2015;169(1-3):458–63.

19 Folch J, Lees M, Sloane Stanley GH. A simple method for the isolation and purification of total lipides from animal tissues. J Biol Chem. 1957;226(1):497–509.

20 Simon-Manso Y, Lowenthal MS, Kilpatrick LE, Sampson ML, Telu KH, Rudnick PA, et al. Metabolite profiling of a NIST Standard Reference Material for human plasma (SRM 1950): GC-MS, LC-MS, NMR, and clinical laboratory analyses, libraries, and web-based resources. Anal Chem. 2013;85(24):11725–31.

21 Fahy E, Sud M, Cotter D, Subramaniam S. LIPID MAPS online tools for lipid research. Nucleic Acids Res. 2007;35(Web Server issue):W606–12.

22 Pluskal T, Castillo S, Villar-Briones A, Oresic M. MZmine 2: modular framework for processing, visualizing, and analyzing mass spectrometry-based molecular profile data. BMC Bioinformatics. 2010;11:395.

23 R Development Core Team. (R Foundation for Statistical Computing, Vienna, 2018).

24 Scrucca L, Fop M, Murphy TB, Raftery AE. mclust 5: Clustering, Classification and Density Estimation Using Gaussian Finite Mixture Models. The R journal. 2016;8(1):289–317.

25 Davison J, O’Gorman A, Brennan L, Cotter DR. A systematic review of metabolite biomarkers of schizophrenia. Schizophr Res. 2018;195:32–50.

26 Westerbacka J, Kotronen A, Fielding BA, Wahren J, Hodson L, Perttila J, et al. Splanchnic balance of free fatty acids, endocannabinoids, and lipids in subjects with nonalcoholic fatty liver disease. Gastroenterology. 2010;139(6):1961–71 e1.

27 Kotronen A, Seppänen-Laakso T, Westerbacka J, Kiviluoto T, Arola JT, Ruskeepää A-L, et al. Hepatic SCD1 activity and diacylglycerol but not ceramide concentrations are increased in the non-alcoholic human fatty liver. Diabetes. 2009;58:203–08.

28 Kotronen A, Velagapudi VR, Yetukuri L, Westerbacka J, Bergholm R, Ekroos K, et al. Serum saturated fatty acids containing triacylglycerols are better markers of insulin resistance than total serum triacylglycerol concentrations. Diabetologia. 2009;52(4):684–90.

29 Oresic M, Hyotylainen T, Kotronen A, Gopalacharyulu P, Nygren H, Arola J, et al. Prediction of non-alcoholic fatty-liver disease and liver fat content by serum molecular lipids. Diabetologia. 2013;56(10):2266–74.

30 Barr J, Caballeria J, Martinez-Arranz I, Dominguez-Diez A, Alonso C, Muntane J, et al. Obesity-dependent metabolic signatures associated with nonalcoholic fatty liver disease progression. J Proteome Res. 2012;11(4):2521–32.

31 Suvitaival T, Bondia-Pons I, Yetukuri L, Poho P, Nolan JJ, Hyotylainen T, et al. Lipidome as a predictive tool in progression to type 2 diabetes in Finnish men. Metabolism. 2018;78:1–12.

32 Rhee EP, Cheng S, Larson MG, Walford GA, Lewis GD, McCabe E, et al. Lipid profiling identifies a triacylglycerol signature of insulin resistance and improves diabetes prediction in humans. J Clin Invest. 2011;121(4):1402–11.

33 Newcomer JW. Antipsychotic medications: metabolic and cardiovascular risk. J Clin Psychiatry. 2007;68 Suppl 4:8–13.

34 Basile VS, Masellis M, McIntyre RS, Meltzer HY, Lieberman JA, Kennedy JL. Genetic dissection of atypical antipsychotic-induced weight gain: novel preliminary data on the pharmacogenetic puzzle. J Clin Psychiatry. 2001;62 Suppl 23:45–66.

35 Theisen FM, Gebhardt S, Haberhausen M, Heinzel-Gutenbrunner M, Wehmeier PM, Krieg JC, et al. Clozapine-induced weight gain: a study in monozygotic twins and same-sex sib pairs. Psychiatr Genet. 2005;15(4):285–9.

36 Pillinger T, McCutcheon RA, Vano L, Mizuno Y, Arumuham A, Hindley G, et al. Comparative effects of 18 antipsychotics on metabolic function in patients with schizophrenia, predictors of metabolic dysregulation, and association with psychopathology: a systematic review and network meta-analysis. Lancet Psychiatry. 2020;7(1):64–77.

37 Carr CN, Lopchuk S, Beckman ME, Baugh TB. Evaluation of the use of low-dose quetiapine and the risk of metabolic consequences: A retrospective review. Ment Health Clin. 2016;6(6):308–13.

38 Borgan F, Laurikainen H, Veronese M, Marques TR, Haaparanta-Solin M, Solin O, et al. In Vivo Availability of Cannabinoid 1 Receptor Levels in Patients With First-Episode Psychosis. JAMA Psychiatry. 2019.

39 Dickens AM, Borgan F, Laurikainen H, Lamichhane S, Marques T, Rönkkö T, et al. Links between central CB1-receptor availability and peripheral endocannabinoids in patients with first episode psychosis. bioRxiv. 2019:664086.

40 Lau BK, Cota D, Cristino L, Borgland SL. Endocannabinoid modulation of homeostatic and non-homeostatic feeding circuits. Neuropharmacology. 2017;124:38–51.

41 Silvestri C, Di Marzo V. The endocannabinoid system in energy homeostasis and the etiopathology of metabolic disorders. Cell Metab. 2013;17(4):475–90.

